# Freezing Prediction Horizon: Quantifying Advanced Warning for Predicting Freezing of Gait in Parkinson’s Disease

**DOI:** 10.64898/2026.01.21.26344382

**Authors:** Mengcong Li, Bohan Shi, Arthur Tay, W.L. Au, Dawn M.L. Tan, Nicole S.Y. Chia, Shih-Cheng Yen

**Author notes:** These authors contributed equally to this work (Mengcong Li, Bohan Shi).

## Abstract

Freezing of gait (FoG) prediction is clinically meaningful only when warnings arrive sufficiently early for subsequent action. Therefore, we adopt a Freezing Prediction Horizon (FPH) evaluation that reports prediction performance as a function of the warning horizon before onset, making the lead-time versus reliability trade-off explicit. Within this protocol, we develop a Transformer-based predictor with a progressive self-paced learning strategy and evaluate it on a 55-patient clinical dataset and two public datasets. The horizon–performance curves show that Macro-F1 remains stable up to approximately 2.5 seconds before FoG onset in our dataset, after which a gradual decline is observed. This horizon-based characterization replaces single, fixed ahead-of-onset windows with a continuous method that summarizes achievable advanced time at specified accuracy levels. In this way, it offers a principled basis for setting targets in real-time implementations-linking algorithmic early-warning capacity to the lead times that practical systems may require-while remaining compatible with conventional metrics. By centering evaluation on FPH, this study clarifies how far in advance FoG can be predicted with confidence, and it positions horizon-based assessment as a reproducible complement to standard reporting for future work on deployable FoG prediction. Ultimately, quantifying advance warning is a prerequisite for prevention-oriented use, by indicating whether sufficient time can be reserved for cueing prior to onset.

## I. INTRODUCTION

Parkinson’s disease (PD) is a progressive neurodegenerative disorder that affects over ten million people worldwide and severely impairs motor control [1–3]. Among its motor complications, freezing of gait (FoG) is particularly disabling. It emerges with disease progression and often becomes prevalent in advanced stages [1]. Clinically, FoG manifests as a brief, involuntary halt of movement despite the intention to walk [4] and is strongly associated with falls, injuries, and loss of independence [5, 6]. The unpredictability of FoG episodes also induces fear of walking and complicates clinical assessment, as short in-clinic tests rarely capture the breadth of real-life events [7].

Current management strategies combine pharmacological, surgical, and rehabilitative approaches [2, 8–10]. Among them, external cueing - providing rhythmic auditory, visual, or vibrotactile stimuli - has proven effective in alleviating ongoing freezing by compensating for impaired internal rhythmicity [11–14]. However, continuous cueing is impractical for daily use and often causes fatigue or habituation, while reactive cueing delivered after a freeze has begun arrives too late to prevent near-falls. These limitations motivate the need for on-demand assistance that activates only when a freeze is imminent. Such an approach requires advance warning-a system capable of predicting FoG early enough for the patient or device to respond safely.

With the emergence of wearable inertial measurement units (IMUs), continuous motion monitoring outside clinical settings has become feasible. IMUs provide rich temporal data that capture subtle gait irregularities and have enabled numerous successful studies on FoG detection, where the goal is to recognize a freezing episode once it has started [15–17]. Building on these advances, researchers have begun exploring FoG prediction, which aims to identify pre-freezing patterns several seconds before onset. Deep learning methods trained on sliding windows of IMU data have achieved encouraging numerical results [18–20].

Despite this progress, current prediction pipelines face key limitations. Most studies typically pool over sliding windows across the entire test stream after defining a fixed pre-onset labeling region (e.g., all samples within 2–5 seconds before onset labeled as pre-FoG). Pooling windows near the onset with those further away obscures how reliability varies with the evaluation horizon (the advance time at which performance is assessed). Consequently, aggregated metrics do not directly reveal how early a model remains dependable, which is a key requirement for practical use, nor do they expose the underlying lead-time–reliability trade-off.

To make this requirement explicit, we use a Freezing Prediction Horizon (FPH) evaluation, which is adapted from seizure prediction [21]. Concretely, we keep the same labeling protocol but condition evaluation on the evaluation horizon *τ*, reporting a horizon–performance profile as *τ* increases. This characterization replaces single-pooled scores with an interpretable summary of achievable advance time at specified accuracy levels, enabling comparisons that are directly relevant to early-warning utility. Therefore, it frames prediction as a prevention-oriented problem: advance warning must be long enough to enable timely, on-demand assistance before freezing occurs.

Building upon this framework, this study makes three key contributions. First, we formalize FPH as a reproducible and leakage-resistant evaluation protocol that quantifies FoG prediction reliability as a function of required lead time, providing a clinically interpretable measure of usable warning duration. Second, we develop a Transformer-based predictor with a progressive self-paced learning strategy, coupled with Bayesian optimization to jointly tune architectural and preprocessing parameters, achieving stable and well-calibrated performance under class imbalance. Finally, we evaluate the proposed framework on a 55-patient clinical dataset and validate it on two public benchmarks (Daphnet [22] and BXHC [23]), demonstrating consistent trends in the horizon–performance curve - maintaining stable Macro-F1 up to approximately 2 s before FoG onset, thereby establishing a reproducible reference for future studies on deployable prediction.

## II. Related Works

Research on freezing of gait prediction has evolved substantially with the rise of wearable inertial measurement units (IMUs). Early studies relied on hand-crafted features derived from accelerometer and gyroscope streams, combined with traditional classifiers. Mazilu et al. [24] demonstrated that statistical and principal-component features from ankle-mounted IMUs could distinguish pre-freezing windows on the Daphnet dataset using decision trees. Palmerini et al. [25] employed cross-correlation and turning-angle measures to capture impaired gait dynamics, applying linear discriminant analysis on CuPiD. Arami et al. [27] introduced time–frequency representations and support vector machines, while Zhang [28] and Borzì [29] modeled step-level asymmetry and spectral energy changes with decision-tree and SVM classifiers. Other variants incorporated entropy- or wavelet-based descriptors [26]. These works collectively established the feasibility of anticipating FoG from lower-limb motion, yet they required extensive feature design and often generalized poorly across tasks or datasets.

The subsequent stage coincided with the adoption of end-to-end deep learning. Convolutional and recurrent architectures began to replace feature pipelines by learning directly from raw IMU sequences. Torvi et al. [**?**] introduced LSTM networks to model temporal dependencies between gait cycles, outperforming manual features on Daphnet. Elziaat et al. [30] transformed tri-axial accelerometer signals into spectrograms for CNN feature extraction, while Naghavi and Wade [31] proposed a multi-scale convolutional backbone combined with transfer learning to stabilize training on small proprietary cohorts. With improved data availability and GPU computing, hybrid and attention-based encoders further advanced the field. Huang et al. [32] implemented a self-attention Transformer that aggregated stride-level dependencies over several seconds on the BXHC dataset. Sun et al. [18] combined manually selected gait descriptors with a deep ResNeXT backbone, balancing interpretability and accuracy, and Wang et al. [33] introduced a multi-channel temporal network jointly modeling accelerometer and gyroscope streams. Xia et al. [20] applied self-supervised contrastive pretraining with Transformers, achieving the strongest published performance on Daphnet and BXHC. As shown in Table I, across these efforts, deep models consistently improved performance compared with classical baselines, confirming the utility of learned temporal representations while reducing dependence on expert-crafted features.

**TABLE I:** Survey of representative FoG prediction methods, summarizing dataset, cohort size (and number of freezers), window configuration, pre-FoG region, feature design, and learning approach. Subject counts in the “Subjects (Freezers)” column refer to the dataset as released, and several studies evaluate on subsets or proprietary cohort. “Window Size (Overlap)” reports the analysis window length and the fraction of overlap between adjacent windows. “Pre-FoG Region” denotes the lead time before FoG onset used to label pre-FoG samples. Feature abbreviations include FI (Freezing Index), SampEn (Sample Entropy), and CWT (Continuous Wavelet Transform).

| Work | Dataset | Subjects (Freezers) | Window Size (Overlap) | Pre-FoG Region | Features | Method |
| --- | --- | --- | --- | --- | --- | --- |
| Mazilu, 2013 [24] | Daphnet | 10 (8) | 1 s (25%) | 3 s | PCA-based unsupervised features | Decision Tree (ML) |
| Palmerini, 2017 [25] | CuPiD | 18 (11) | 2 s (0%) | 2 s | · Left-right cross correlation · Turning degrees · FI | LDA (ML) |
| Naghavi & Wade [26] | Daphnet | 10 (8) | 2 s (75%) | 2 s | · SampEn · FI · CWT | Threshold-based classification with statistical inference |
| Arami [27] | Daphnet | 10 (8) | 4 s (87.5%) | 3.84 s | Time–frequency features | SVM-RBF (ML) |
| Zhang [28] | Proprietary | 12 | 2 steps (50%) | 6 steps | Step-based impaired gait features | Decision Tree (ML) |
| Borzi [29] | Proprietary | 11 | 2 s (0%) | 2 s | Time-frequency step features | SVM (ML) |
| Elziaat [30] | Daphnet | 10 (8) | 1 s (50%) | 2 s | Spectrogram-CNN features | RF, SVM, etc. (ML) |
| Naghavi & Wade [31] | Proprietary | 7 | 2 s (12.5%) | 1 s | Raw IMU features | Transfer Learning with CNN (DL) |
| Huang [32] | BXHC | 12 (11) | 3 s (83.3%) | 5 s | Raw IMU features | Self-attention Transformer (DL) |
| Sun [18] | Daphnet | 10 (8) | 1 s (50%) | 5 s | Manually selected gait features | ResNeXT (DL) |
| Wang [33] | Daphnet | 10 (8) | 2 s (75%) | 2 s | Raw IMU features | Multi-channel time-series neural network (DL) |
| Xia [20] | Daphnet | 10 (8) | 1 s (50%) | 5 s | Raw IMU features | Self-supervised contrastive learning with Transformer (DL) |
| Xia [20] | BXHC | 12 (10) | 1 s (50%) | 5 s | Raw IMU features | Self-supervised contrastive learning with Transformer (DL) |

In these works, prediction tasks are generally defined in two distinct ways that differ in temporal granularity and evaluation focus. The window-level formulation divides continuous recordings into overlapping fixed-length segments labeled according to gait state, typically using a three-class scheme of normal gait, pre-FoG, and FoG [18, 20, 24, 26, 27, 30]. Some studies simplify this structure by merging categories to reduce class imbalance [27, 29, 34], while others expand it to include post-FoG recovery, as in Wang et al. [33]. This formulation treats prediction as a window-based classification problem and remains the most common design for short-horizon forecasting.

The episode-level formulation, by contrast, evaluates prediction at the scale of entire freezing episodes. Rather than assigning labels to every window, it quantifies whether an impending episode is successfully anticipated and how much time separates the first alarm from the onset. Huang et al. [32] exemplified this paradigm by defining a time-margin metric on the BXHC dataset to measure the effective early-warning interval. Each formulation offers distinct advantages. Window-level methods provide fine-grained temporal supervision and can train on relatively small datasets, whereas episode-level analyses align more closely with the operational goal of warning. Yet both typically summarize results as aggregated accuracy or F1 scores, masking how performance evolves with increasing lead time before onset.

Despite rapid algorithmic progress, IMU-based FoG prediction still faces constraints that arise from both the data regime and the evaluation regime, which together shape how results should be interpreted. On the data side, most public and proprietary cohorts include only a few dozen freezers and relatively few freezing episodes, limiting statistical power and encouraging models to fit subject-specific idiosyncrasies [22, 23]. A further consequence is severe class imbalance that pre-FoG segments are short and scarce relative to normal gait and FoG, and across window-level studies, they typically yield the weakest per-class scores even when headline metrics appear high, reflecting intrinsic ambiguity and temporal heterogeneity [**?**, 18, 20, 26, 27, 30, 33].

On the evaluation side, most reports compute a single pooled metric over all sliding windows in the test stream, which mixes onset-adjacent windows with those farther in advance. This aggregation obscures the relationship between achievable lead time and reliability, and can permit subtle temporal leakage when pre-onset evidence overlaps with labeled FoG segments [18, 20, 24, 27, 30, 33]. Episode-level analyses that quantify an early-warning margin begin to address clinical relevance [32], but they are sensitive to alarm-definition details and remain less common given the small number of episodes per subject. Taken together, and compounded by cross-paper differences in windowing, sampling rates, sensor configurations, and post-processing, these factors undermine comparability across studies and weaken the link between reported scores and actionable early warning.

## III. Materials and Methods

### A. Dataset

Three Freezing of Gait (FoG) datasets were used in this study, as summarised in Table II.

**TABLE II:** Summary of publicly available and clinical FoG datasets used in this study. The table lists the number of participants (and freezers), sensor modalities, placement location, and sampling frequency (*f*_*s*_) for each dataset. These specifications highlight the diversity of sensing setups and acquisition protocols considered in the cross-dataset evaluation.

| Dataset | Subjects (Freezers) | Sensor Types | Placement | $f_s$ (Hz) |
| --- | --- | --- | --- | --- |
| Daphnet [22] | 10 (8) | Accelerometer | Shank Thigh Trunk | 64 |
| BXHC [23] | 12 (10) | Accelerometer Gyroscope | Waist Shanks | 500 |
| Our Dataset | 63 (55) | Accelerometer Gyroscope Magnetometer | Ankles | 50 |

The Daphnet dataset [22] contains nine accelerometer traces per trial-three each on shank, thigh, and trunk - from ten people with Parkinson’s disease, eight of whom are classified as freezers. Signals were sampled at 64 Hz and annotated frame-wise for FoG by clinical experts.

The BXHC dataset [23] includes multi-modal recordings collected at Beijing Xuanwu Hospital center. In the present work only the inertial channels are used: tri-axial accelerometer and gyroscope signals from the waist and both shanks, sampled at 500 Hz. Twelve Parkinson’s disease participants (ten freezers) performed continuous walking tasks, manually tagged in synchronized video streams.

For our dataset, ethical approval was obtained from the SingHealth Centralised Institutional Review Board (CIRB Ref: 2016/2743) on 28 September 2016. Data collection occurred at two Singapore National Neurological Institute clinics from September 2016 to August 2017. Sixty-three Parkinson’s disease subjects were recruited through convenience sampling.

Each subject was instructed to perform the 7-meter Timed Up-and-Go (7mTUG) test thrice, with an additional free walk test for those not exhibiting FoG during 7mTUG. In-house developed IMUs were attached to the subjects’ ankles and the seventh cervical (C7) vertebra. Each IMU, comprising an accelerometer, gyroscope, and magnetometer, sampled data at 50 Hz and transmitted it via Bluetooth 4.0. The neck IMU was primarily used to analyse the subject’s sitting and standing posture and stability during the walk. The data from the neck IMU was excluded from the FoG prediction model due to its capture of both lower and upper body movements, which could introduce unnecessary noise and redundancy.

All tests were video recorded to ensure accurate labeling of FoG episodes. After reviewing the video and sensor data, we found that only fifty-five out of the sixty-three subjects manifested at least one FoG episode during the tests. To focus on FoG and the brief transitionary window preceding it, we used data exclusively from subjects who experienced freezing during the clinic visits. A total of 481 independent FoG episodes were captured across the two ankle IMUs, allowing our prediction model to detect any pre-FoG episodes captured by either IMU without the need for additional data processing to synchronize data from different limbs. This design also increases the likelihood of early FoG prediction because FoG may occur slightly earlier in one limb than the other. Further details about the dataset are available in our previous work [17, 35].

### B. Data Preprocessing

As illustrated in Fig. 1A, before training the model, the raw inertial measurement unit (IMU) signals underwent a series of preprocessing steps to ensure data consistency and reduce noise. First, we applied normalization across all sensor channels to unify the magnitude of accelerometer, gyroscope, and magnetometer readings, ensuring that different input features remain within a comparable numerical range. Additionally, to remove high-frequency noise and preserve critical gait-related information, a low-pass filter was employed. Following this, the time-series data were segmented into fixed-size sliding windows with a predefined overlap ratio, facilitating sequential input for the deep learning model.

**Fig. 1:**
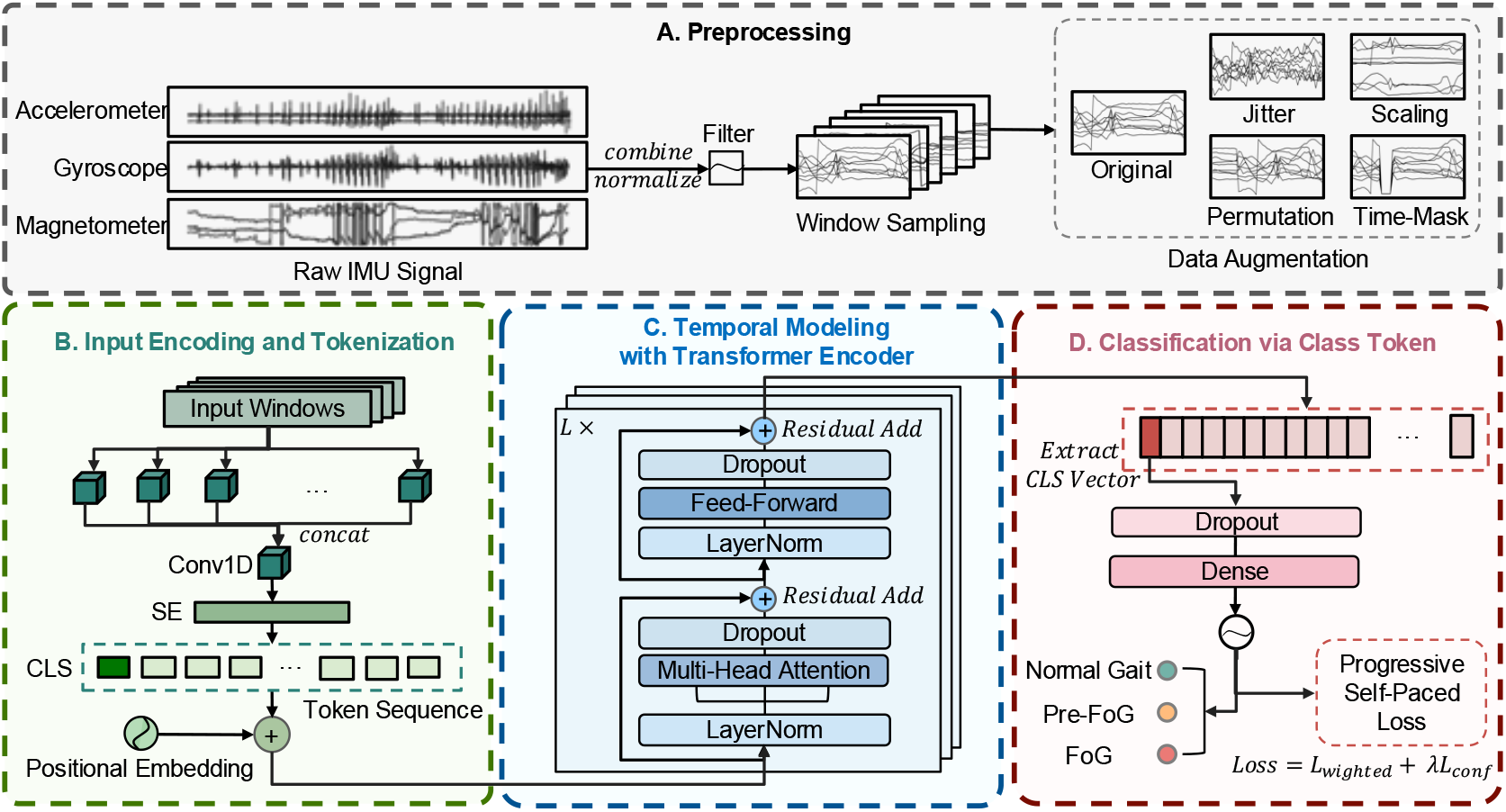
Overview of the proposed FoG prediction framework. **A. Preprocessing:** Raw IMU signals (accelerometer, gyroscope, magnetometer) are combined, normalized and low-pass filtered, then segmented into overlapping windows. Data augmentation (jittering, magnitude scaling, segment permutation, time masking, mixup) is applied on each window. **B. Input Encoding and Tokenization:** A multi-scale convolutional stem captures local temporal features, followed by squeeze-and-excitation channel attention. A learnable class token and positional embeddings are prepended to form the transformer input sequence. **C. Temporal Modeling with Transformer Encoder:** Stacked Pre-Norm Transformer blocks perform multi-head self-attention and feed-forward transformations with residual connections to model long-range dependencies across the gait window. **D. Classification via Class Token:** The final class token is extracted, passed through a dropout and dense layer, and supervised with a progressive self-paced loss combining confidence-weighted cross-entropy and confidence regularization.

A significant challenge in freezing of gait (FoG) prediction is the inherent class imbalance, where non-FoG samples vastly outnumber FoG and pre-FoG instances. To mitigate this imbalance, we incorporated various data augmentation techniques. Spatial-temporal augmentation includes transformations that directly modify the structure of time-series gait data while preserving its underlying physiological characteristics. Specifically, we applied random jittering, magnitude scaling, segment permutation, and time masking. The random jittering operation introduced small random perturbations to the input signals by adding Gaussian noise, simulating sensor instability and natural variation. Magnitude scaling applied random channelwise scaling factors to simulate changes in signal amplitude due to varying sensor sensitivity or user-specific movement intensity. Segment permutation shuffled predefined temporal segments within a sample, helping the model learn robust temporal representations. Temporal masking randomly zeroed out continuous portions of the signal, encouraging the model to remain resilient to missing or dropped sensor data.

To ensure that augmentation does not introduce biases in model evaluation, all augmentation techniques were applied only to the training set. These preprocessing and augmentation strategies collectively improved the model’s ability to handle noisy, imbalanced, and real-world FoG prediction scenarios, thereby enhancing the reliability of its predictions.

### C. Model Architecture

The proposed model is designed to anticipate Freezing of Gait (FoG) events by analyzing short-term and long-range dependencies in multichannel IMU data. As shown in Figure 1 B-D, it consists of three main stages: multi-scale convolutional encoding with squeeze–excitation and class token injection, temporal modeling via stacked Transformer encoders, and final classification through the class token.

#### 1) Input Encoding and Tokenization

The goal of this stage is to transform raw IMU signals into a representation that captures local temporal features while aligning with the Transformer model’s input format. Given a time-series window *x* = [*x*_1_, *x*_2_, …, *x*_*T*_] ∈ ℝ^*T*×*D*^, where *T* is the number of time steps and *D* is the number of sensor channels, we first apply a multi-scale convolutional stem to extract localized motion patterns such as gait oscillations and step cycles. Each time step is passed through multiple parallel 1-D convolutions followed by batch normalization and GELU:

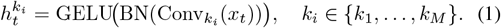

The branch outputs are concatenated, channel-compressed with a 1 × 1 convolution, and re-weighted by a squeeze–excitation (SE) block:

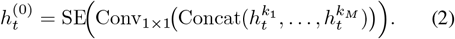

This captures small-scale dynamics that often precede FoG onset, especially subtle reductions in movement amplitude during pre-FoG phases.

If the stem output width differs from the Transformer dimension *d*, a 1 × 1 point-wise convolution projects the features:

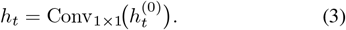

The resulting tokens [*h*_1_, …, *h*_*T*_] are prepended with a learnable class token *x*_cls_ ∈ ℝ^*d*^ and augmented by positional embeddings:

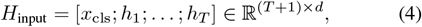

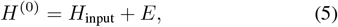

Here, *E* ∈ *ℝ* ^(*T* +1)×*d*^ encodes relative position, which is critical for temporal modeling where the order of gait phases matters.

#### 2) Temporal Modeling with Transformer Encoder

To model long-range dependencies within each gait window, the encoded sequence *H*^(0)^ ∈ ℝ ^(*T* +1)×*d*^ is passed through a stack of *L* Transformer encoder layers. Each layer *𝓁* consists of a multi-head self-attention (MHSA) module and a feed-forward network (FFN), both preceded by layer normalization and followed by residual connections:

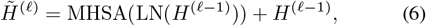

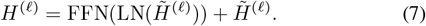

The MHSA module applies scaled dot-product attention over multiple heads and enables tokens to attend to all positions in the sequence. Outputs from all heads are concatenated and projected back to the model dimension. The FFN is applied independently to each token and consists of two linear layers with a GELU activation:

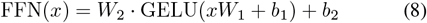

where 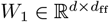 and 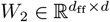.

This architecture enables flexible integration of both local and global temporal cues. Importantly, the class token is updated at each layer via self-attention, progressively aggregating sequence-level context. This is especially valuable for anticipating pre-FoG episodes, where predictive cues may be sparse and temporally diffuse.

#### 3) Classification via Class Token

After the final Transformer layer, we extract the output of the class token:

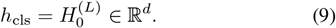

A linear classifier then maps this vector to class probabilities:

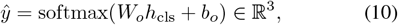

where *W*_*o*_ ∈ ℝ^3×*d*^ and *b*_*o*_ ∈ ℝ^3^ are learnable parameters. The predicted vector *ŷ* represents the probability distribution over three classes: normal gait, pre-FoG, and FoG. This classification strategy leverages the global aggregation ability of the class token (CLS token). By summarizing information across all time steps, the token enables the model to anticipate upcoming FoG events based on distributed and subtle gait anomalies.

This model architecture is tailored for the anticipatory nature of FoG prediction: the convolutional encoder captures short-term gait fluctuations, the Transformer models temporal evolution, and the class token aggregates holistic sequence-level information. This integration enables precise and early detection of pre-FoG states which is critical for timely clinical intervention.

### D. Progressive Training Strategy

In the context of Freezing of Gait (FoG) prediction, severe class imbalance and label ambiguity - especially for pre-FoG samples - make stable optimization challenging. To address these issues, we adopt a Progressive Self-Paced Learning strategy that adjusts each sample’s contribution to the total loss according to its predicted confidence. Unlike conventional progressive learning that gradually enlarges the network or dataset, our method is progressive in the sense that sample weights evolve progressively with training, following the self-paced learning principle. Specifically, the framework combines a confidence-weighted classification loss and a confidence-regularization term, allowing the model to first focus on reliable samples and then gradually incorporate more uncertain ones.

#### 1) Confidence-Weighted Loss Function

Let **x**_*i*_ ∈ ℝ^*T* ×*D*^ denote the *i*-th input time-series window, and **y**_*i*_ ∈ {0, 1}^*C*^ its corresponding one-hot label, where *C* = 3 denotes the target classes: normal gait, pre-FoG, and FoG. The model outputs a predicted probability distribution **Ŷ**_*i*_ = *f*_*θ*_(**x**_*i*_) ∈ [0, 1]^*C*^ through a softmax function. We define the confidence of the prediction as:

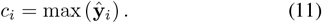

A piecewise weighting function *w*(*c*_*i*_) is used to modulate the loss:

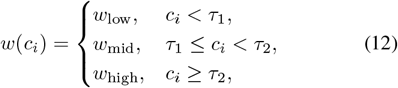

where (*τ*_1_, *τ*_2_) = (0.6, 0.8) and (*w*_low_, *w*_mid_, *w*_high_) = (0.5, 0.7, 1.0). The classification loss is then defined as:

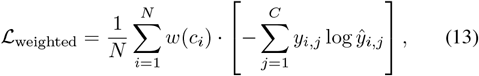

which effectively down-weights uncertain or noisy samples early in training, and gradually increases their contribution as the model becomes more confident. This mechanism is particularly valuable for pre-FoG classification, where subtle transitions and label ambiguity often result in low-confidence predictions at the beginning of training.

#### 2) Confidence Regularization

To further enforce prediction calibration, we introduce a regularization term that softly constrains each sample’s confidence toward a target level *c*\* ∈ (0, 1). This term is defined as:

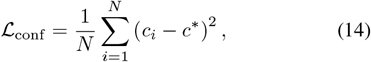

where *c*_*i*_ is the maximum probability in **Ŷ**_*i*_. We set *c*\* = 0.8. This regularization encourages well-calibrated predictions: it discourages premature overconfidence in ambiguous transitions (e.g., early pre-FoG stages) and ensures stable learning across all classes, especially for underrepresented FoG and pre-FoG segments.

#### 3) Training Procedure

The total loss combines the classification and regularization components:

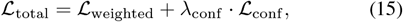

where *λ*_conf_ balances the influence of the confidence regularization.

Training is performed using the AdamW optimizer with a warm-up cosine decay scheduler. Early stopping is applied based on the validation Macro F1 score, and we retain the checkpoint that yields the highest validation Macro F1 score.

Overall, this Progressive Self-Paced Learning strategy enhances generalization and robustness, particularly for the ambiguous pre-FoG class, which is inherently difficult to learn due to temporal fuzziness and class imbalance. By progressively increasing the influence of harder, low-confidence samples while maintaining calibrated predictions, the model learns in a self-regulated manner and achieves more reliable early forecasts of impending FoG episodes.

### E. Freezing Prediction Horizon (FPH) Protocol

The Freezing Prediction Horizon (FPH) protocol quantifies how early a forthcoming FoG episode can be predicted in a way that remains reliable and clinically useful. Inspired by the Seizure Prediction Horizon in epilepsy, FPH withholds onset-proximal data during evaluation so that any reported performance corresponds to decisions made sufficiently in advance for potential intervention [19, 21].

In practice, FPH imposes a simple test-time rule around each annotated onset that for a chosen horizon (denoted *τ* seconds), windows whose centers fall within the last *τ* seconds before the onset are excluded from evaluation. When *τ* = 0, only the FoG windows themselves are withheld, and larger *τ* progressively removes windows closer to onset, forcing predictions to rely on gait information further in advance. Figure 2 illustrates this masking of onset-proximal segments.

**Fig. 2:**
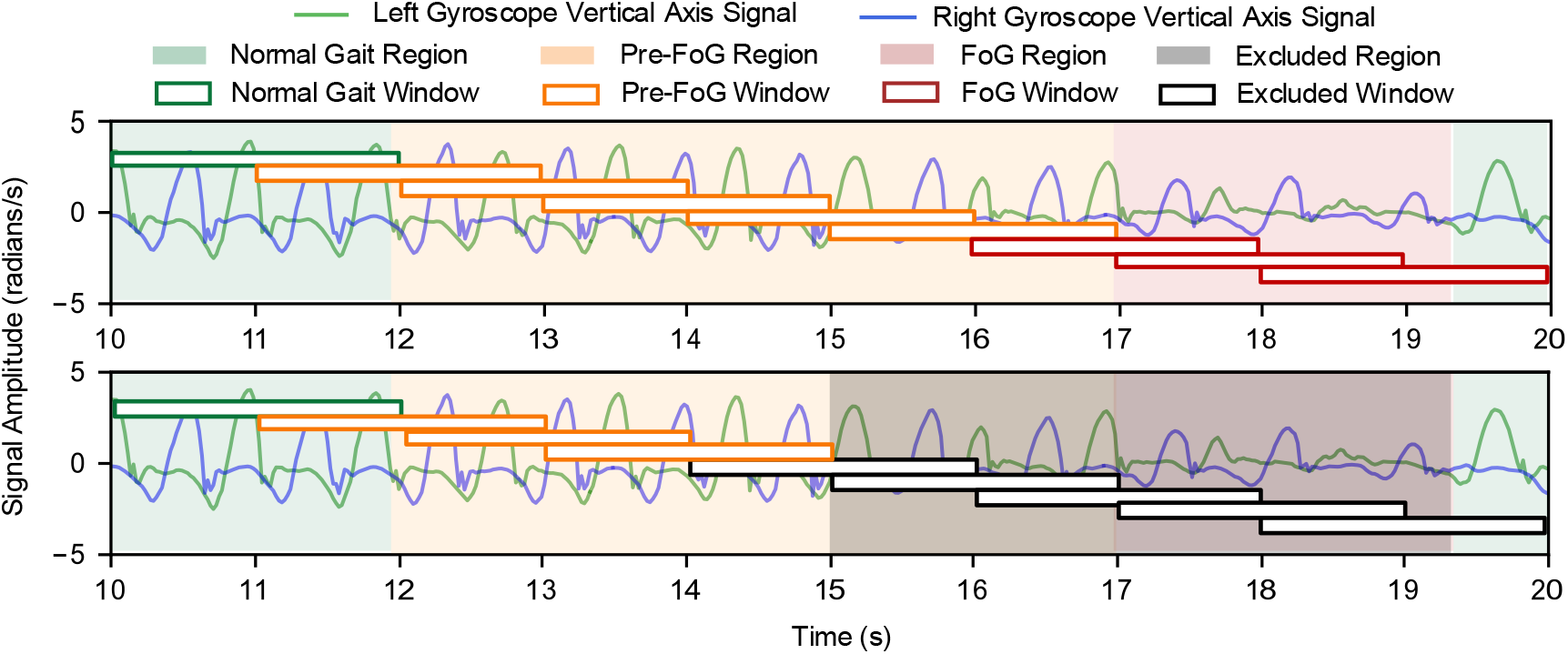
Illustration of the Freezing Prediction Horizon (FPH) protocol. The top panel shows a 10 s signal segmented into Normal gait (green), Pre-FoG (orange), and FoG (red) windows. In this example, the horizon *τ* is set to 2 s. The bottom panel depicts the testing stage: all FoG windows and any window whose center falls within (*t*_0_ − *τ, t*_0_] before a FoG onset are masked and discarded (grey), ensuring that the model receives no information from the interval immediately preceding the FoG event.

Crucially, the horizon constraint is applied only at test time. Training data, label definitions (including any pre-FoG labeling used to build the dataset), and model parameters are fixed. Sweeping *τ* over a range yields a horizon-indexed performance curve under identical training conditions, which isolates genuine forecasting ability from labeling conventions and avoids inflation due to near-onset detection. Because masking is applied per patient and per onset, results remain comparable across horizons while respecting subject-specific timing.

Read as a function of *τ*, FPH transforms early prediction into a quantitative warning time standard. Stability of performance beyond a given *τ* implies headroom for practical pipelines that incur transmission, processing, and device-actuation delays; conversely, performance that degrades before a required *τ* signals insufficient time for preventive cueing. At large horizons valid test windows can become sparse; therefore, we report the number of contributing subjects/windows alongside metrics to make statistical uncertainty explicit. In sum, FPH provides a reproducible and leakage-free link between algorithmic outputs and the question that matters for prevention: how much dependable time is available before freezing occurs.

## VI. Results

All experiments were carried out in Python 3.11.8 with TensorFlow 2.15 on the NUS high-performance computing cluster (Vanda) equipped with NVIDIA Tesla V100 GPUs.

### A. Patient Dependent Evaluation on Our Dataset

Before presenting the main results, we note that the choice of data parameters and model configuration was determined through Bayesian optimization (BO), as detailed in Appendix I. This process identified a 5 s pre-FoG region, a 1 s window size, and an optimized Transformer architecture.

We first evaluated the proposed framework under the PD scenario using stratified 5-fold cross-validation on our clinical dataset comprising 55 subjects, as summarized in Figure 3. Macro F1 and Geo-mean were chosen as the primary metrics, given their balanced assessment of multi-class classification and robustness under severe class imbalance.

**Fig. 3:**
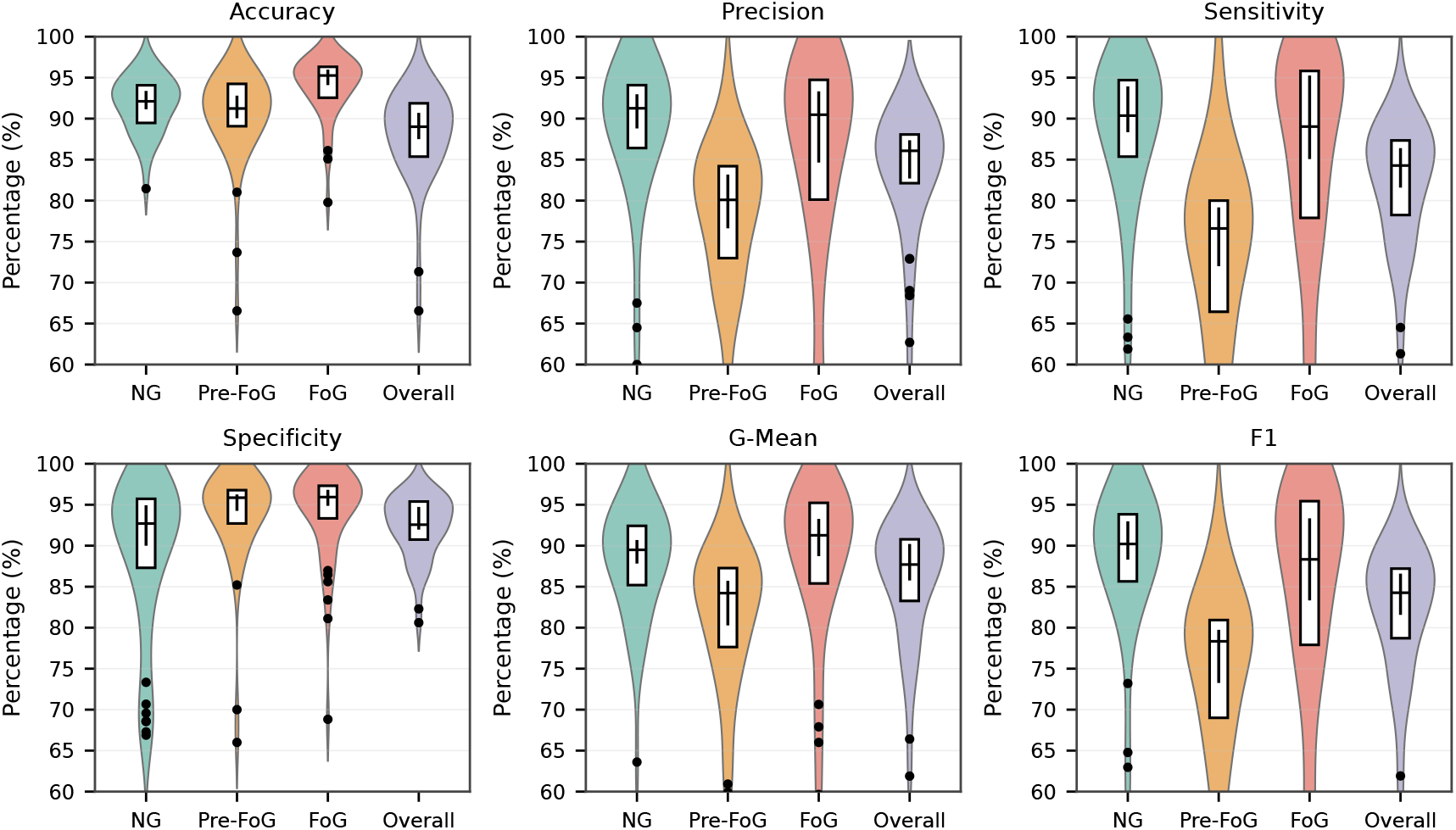
Patient dependent (PD) performance on our clinical dataset visualized using violin plots across 55 subjects. Each panel shows the distribution of Accuracy, Precision, Sensitivity, Specificity, G-Mean, and F1-score for Normal gait (NG), Pre-FoG, FoG, and the overall (Macro) average. The colored violins represent the density of subject-level scores for each class. For each violin, the white box indicates the interquartile range (25th–75th percentile), the horizontal line inside the box marks the median, and the vertical black line shows the bootstrapped 95% confidence interval (CI) of the median across subjects. Black dots outside the box-and-whisker range denote outliers.

Figure 3 presents the full distribution of PD performance across subjects using violin plots. Each violin visualizes the variability and consistency of subject-level scores, with the white dot and vertical bar indicating the mean ± 95% confidence interval (CI). Detailed numerical values are provided in Appendix II (Table V). The model achieved strong performance for normal gait (F1 = 90.21%), while pre-FoG detection remained more challenging (F1 = 75.07%), consistent with its transient and subtle manifestation.

Overall, the framework achieved strong and stable patient-dependent performance. Across the 55 subjects, the Macro F1 averaged 84.32 ± 3.11%, while the median Macro F1 was 85.22% (95% CI: [81.51, 86.88]), indicating both high overall accuracy and low variability across folds. Statistical comparisons across classes (Kruskal–Wallis followed by

Welch–Holm tests) revealed that performance differences were significant for both F1 (*p <* 0.001) and G-Mean (*p <* 0.001). In particular, Pre-FoG exhibited significantly lower F1 scores than both Normal gait and FoG (*p <* 0.001), consistent with its role as the most ambiguous and challenging transitional phase. Effect-size analysis further confirmed substantial separation, with large Cliff’s delta values when comparing Pre-FoG against Normal gait (| *δ*| = 0.68–0.74) and moderate-to-large effects against FoG (|*δ*| = 0.41–0.54). Class-wise specificity consistently exceeded 95% for both pre-FoG and FoG classes, demonstrating that the model effectively minimizes false alarms - a critical requirement for integration into assistive cueing devices.

These results establish a reliable baseline and motivate a deeper analysis of how predictive ability evolves when progressively stricter advance-warning requirements are imposed.

### B. Performance under the FPH Protocol

We next investigated how far in advance reliable prediction can be sustained by applying the Freezing Prediction Horizon (FPH) protocol under the same PD setting. For each horizon *τ*, test windows whose centers fell within (*t*_0_ − *τ, t*_0_] were excluded, and inference was repeated on the remaining data to compute horizon-specific Macro F1 scores. Horizons were varied from 0 to 5 s in increments of 0.5 s.

Figure 4 illustrates the distribution of Macro F1 across horizons. Performance gradually declined as *τ* increased, reflecting the reduced availability of informative pre-onset windows. This trend is expected because windows near the freezing onset contain the strongest predictive cues, which are progressively removed when the horizon lengthens. The decrease was moderate for short horizons but accelerated once *τ* exceeded 2 s, indicating that gait alterations within the final 2 s before onset are the most discriminative for FoG prediction. The lower panel also shows how class composition evolves concurrently: as *τ* grows, pre-FoG samples are increasingly eliminated, leading to higher imbalance and wider variability in results.

**Fig. 4:**
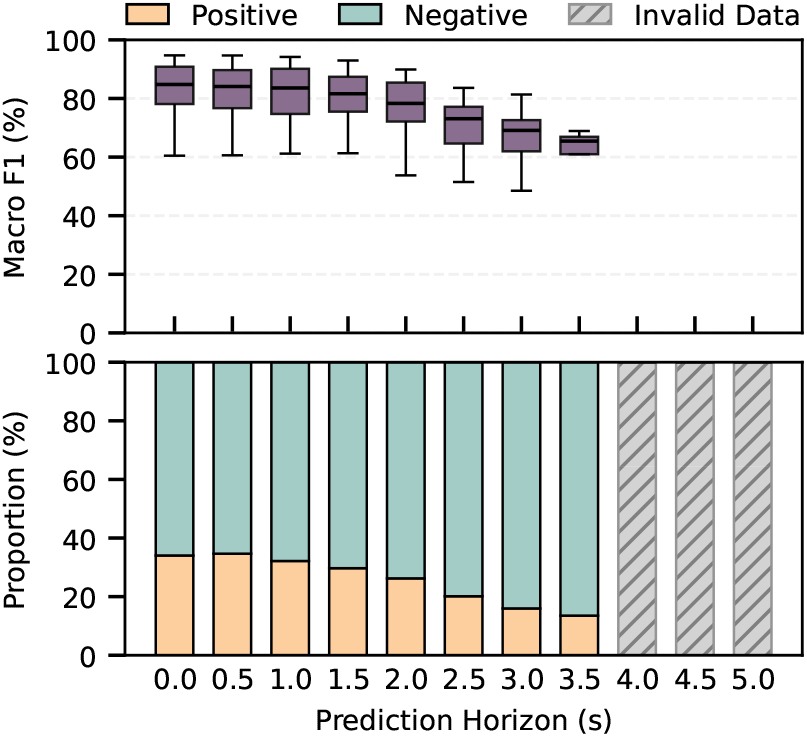
Performance under the Freezing Prediction Horizon (FPH) protocol. Top panel: Boxplots showing the distribution of Macro F1 scores at each horizon *τ* from 0 to 5 s. Bottom panel: Stacked bar plot illustrating the average proportion of pre-FoG (positive) and normal gait (negative) labels remaining in the test set after exclusion. As *τ* increases, the number of pre-FoG samples decreases markedly, leading to greater class imbalance and increased variability in the reported Macro F1. For larger *τ*, some subjects contribute no valid windows, hence fewer samples are available.

To statistically quantify the effect of increasing horizon lengths, we averaged Macro F1 scores across folds within each subject, yielding one subject-level value per horizon *τ*. Because the same subjects appear repeatedly across horizons, the observations are not independent, and their within-subject correlation violates the assumptions of classical oneway ANOVA. We therefore employed a Linear Mixed-Effects Model (LMM) treating horizon *τ* as a fixed effect and subject identity as a random effect to account for repeated measures and unbalanced samples. Post-hoc pairwise comparisons against the baseline (*τ* = 0 s) were adjusted using Holm’s method to control the family-wise error rate (*α* = 0.05). As summarized in Table III, performance remained statistically comparable to the baseline up to *τ* = 2.0 s (*p*_adj_ *>* 0.05), whereas a significant decline emerged from *τ* = 2.5 s onward (*p*_adj_ *<* 0.05). Together, these results suggest a reliable prediction horizon of approximately 2 s, beyond which performance deteriorates sharply.

**TABLE III:** Pairwise comparisons of subject-level Macro F1 between the baseline (*τ* = 0.0 s) and each Freezing Prediction Horizon (FPH) using a Linear Mixed-Effects Model (LMM). Fixed effects were estimated with patient-specific random intercepts. ΔMacro–F1 denotes the estimated difference relative to the baseline; *p*_adj_ are Holm-adjusted values for multiple comparisons. Bold *p*_adj_ indicates significance at *α*=0.05. For *τ* ≥ 3.5 s, insufficient valid samples remained after exclusion, hence comparisons were not computed.

| FPH $\tau$ (s) | ΔMacro F1 | Std. Error | $p$ | $p_{\text{adj}}$ |
| --- | --- | --- | --- | --- |
| 0.5 | -0.0097 | 0.0069 | 0.1589 | 0.3178 |
| 1.0 | -0.0054 | 0.0075 | 0.4749 | 0.4749 |
| 1.5 | -0.0138 | 0.0082 | 0.0929 | 0.3041 |
| 2.0 | -0.0198 | 0.0112 | 0.0760 | 0.3041 |
| **2.5** | -0.0539 | **0.0144** | **0.0002** | **0.0011** |
| **3.0** | -0.0519 | **0.0181** | **0.0041** | **0.0203** |
| 3.5 | - | - | - | - |
| 4.0 | - | - | - | - |
| 4.5 | - | - | - | - |
| 5.0 | - | - | - | - |

Together, these results provide the systematic characterization of an FoG prediction horizon. They demonstrate that our framework not only achieves strong baseline PD performance but also sustains clinically meaningful advance warning, thereby addressing a key gap in prior FoG prediction literature.

### C. Subject-Level Performance Analysis

To investigate the reliability of the proposed method at the individual level, we analyzed the distribution of subject-specific Macro F1 scores at the critical prediction horizon of *τ* = 2.0 s. As shown in Figure 5, the majority of subjects demonstrated high predictability. However, we observed a long-tail distribution where a small subset of patients exhibited suboptimal performance. Using a threshold of 75%, we identified a “Low Performance” group comprising 6 out of 43 subjects (14.0%), while the remaining 37 subjects (86.0%) maintained high prediction accuracy.

**Fig. 5:**
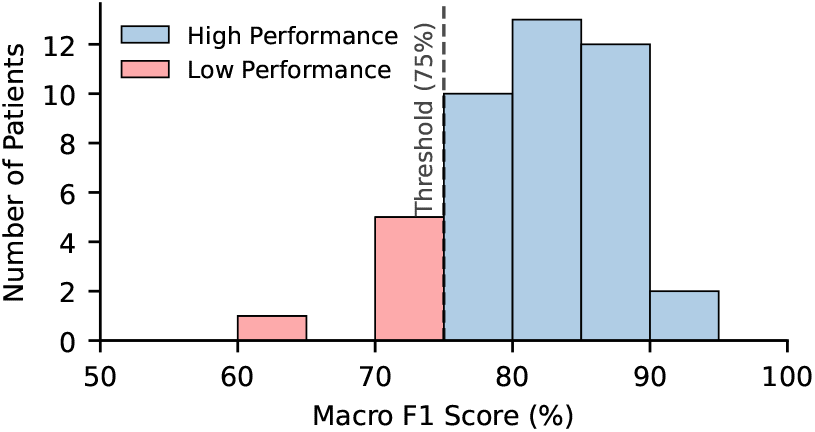
Distribution of subject-level Macro F1 scores at the 2 s Freezing Prediction Horizon (FPH), with subjects grouped according to a 75% performance threshold.

We further examined whether these performance discrepancies were driven by patient demographics or disease severity. We compared the High and Low performance groups across four key clinical metrics: the Freezing of Gait Questionnaire (FOGQ) score, Age, Disease Duration, and Timed Up and Go (TUG) time. The comparative results are presented in Figure 6.

**Fig. 6:**
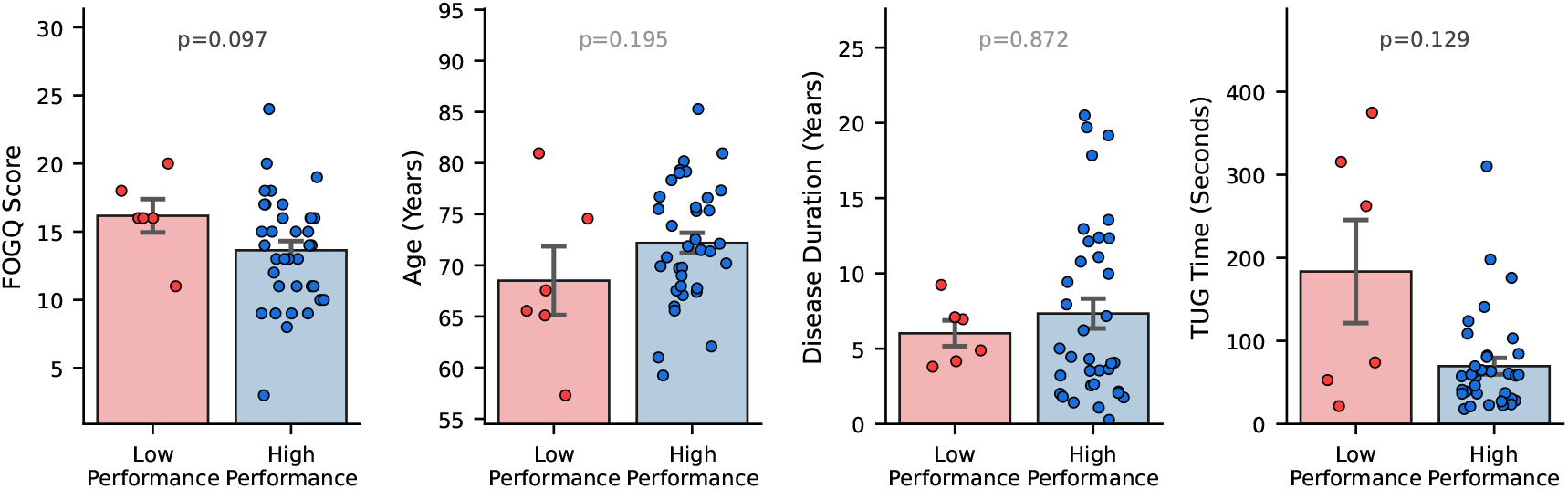
Comparison of clinical variables (FoG-Q score, age, disease duration, and TUG time) between the high- and low-performance groups at the 2 s Freezing Prediction Horizon (FPH).

Statistical analysis revealed no significant differences between the two groups across any of the measured variables (all *p >* 0.05). While the Low Performance group showed a slight trend towards higher FOGQ scores (*p* = 0.097) and longer TUG times (*p* = 0.129), hinting at potentially more severe motor impairment, the absence of significance suggests that disease severity or demographic variables do not systematically explain which subjects are difficult to predict at the 2 s horizon.

Overall, this analysis reinforces that the proposed model maintains stable early-warning performance for the vast majority of subjects at the 2 s horizon. At the same time, the lack of association between clinical characteristics and prediction accuracy suggests that conventional metadata do not capture the factors underlying subject-level variability. This observation is consistent with prior findings that pre-FoG gait changes can manifest differently across individuals, even when clinical severity appears comparable. The small subset of low-performing cases, therefore likely reflects heterogeneous gait dynamics rather than differences in disease stage, highlighting an intrinsic challenge for FoG prediction that is not observable through standard clinical assessments.

### D. Cross-Dataset Patient-Dependent Evaluation

To examine performance beyond our own dataset, we conducted PD experiments on two public datasets - Daphnet [22] and BXHC [23], as summarized in Figure 7. For each dataset, Bayesian optimization (BO) was applied independently following the same search protocol described in Appendix I, jointly tuning data and model parameters to evaluate the adaptability of the proposed framework under heterogeneous conditions.

**Fig. 7:**
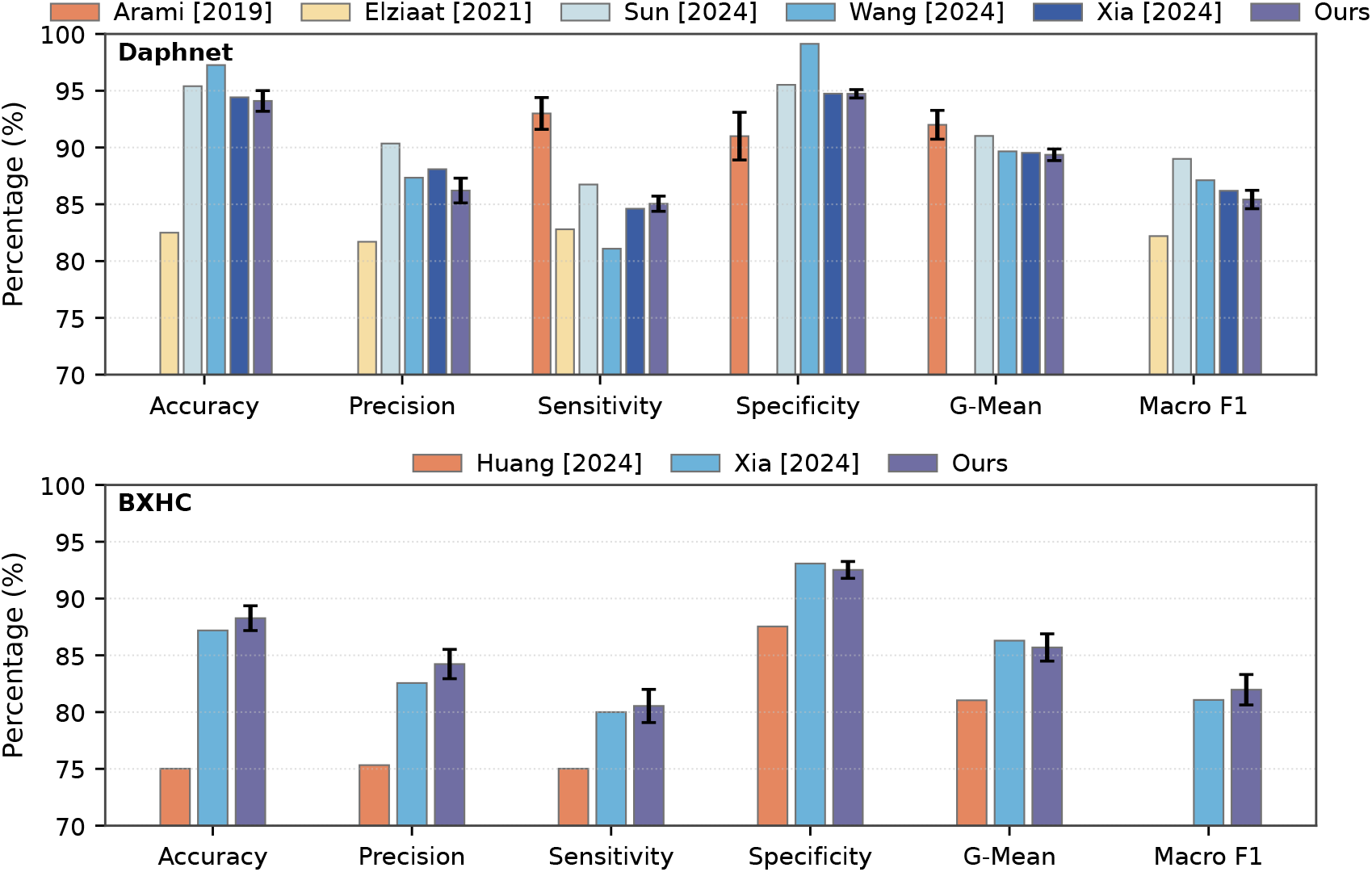
Comparison of patient-dependent (PD) performance across datasets under the Stratified K-Fold evaluation. Each bar represents the mean and standard error (SE) across folds, where the SE for Arami [27] was estimated from the reported standard deviation (SD). Results are shown for the Daphnet (top) and BXHC (bottom) datasets. The proposed model demonstrates competitive accuracy, precision, and sensitivity, while maintaining balanced specificity, G-mean, and Macro F1 across datasets.

Figure 7 summarizes the cross-dataset PD results alongside representative prior work. Detailed numerical values are provided in Appendix II (Table VI). On Daphnet, our framework achieved a Macro F1 of 85.42 ± 0.81% and a Geo-mean of 89.36 ± 0.51%. These values exceed those reported by Elziaat et al. [30], who used CNN-based spectrogram features with classical ML classifiers, highlighting the advantages of deep temporal modelling. Compared with more recent approaches [18, 20], our performance is slightly lower in absolute terms, but remains competitive while using a unified optimization strategy without dataset-specific architectural modifications. The improvements over earlier baselines and the relatively small gap to the latest SOTA confirm that our progressive self-paced learning and Transformer-based framework provides a strong balance between robustness and simplicity.

**TABLE IV:** Search space and optimal values selected via Bayesian Optimization. *R*: real range, *Z*: discrete domain.

| Parameter | Search Space | Optimal |
| --- | --- | --- |
| **Model Parameters** |  |  |
| Dropout Rate | $R \in [0.01, 0.5]$ | 0.25 |
| Learning Rate | $R \in [10^{-4}, 10^{-2}]$ | 0.00095 |
| No. of Attention Heads | $Z \in [2, 8]$ | 2 |
| Hidden Dimension | $Z \in [32, 512]$ | 256 |
| No. of Transformer Blocks | $Z \in [1, 8]$ | 4 |
| FFN Multiplier | $Z \in [1, 10]$ | 7 |
| SE Ratio | $Z \in [4, 16]$ | 8 |
| Stem Channels | $Z \in [64, 256]$ | 64 |
| **Data Parameters** |  |  |
| Pre-FoG Duration (s) | $Z \in \{2, 3, 4, 5, 6\}$ | 5 |
| Window Size (s) | $Z \in \{0.5, 1.0\}$ | 1.0 |

On BXHC, our framework reached a Macro F1 of 81.97 ± 1.34% and a Geo-mean of 85.69 ± 1.20%. These results achieve comparable Macro-F1 to Xia [20], with a slightly lower Geo-mean. Considering that BXHC provides only six sensor channels (waist and a single shank) compared to the nine channels in Daphnet, while also involving a dataset with notable inter-patient variability, the ability to achieve balanced scores across classes is encouraging. In particular, balanced specificity was maintained for both FoG and pre-FoG classes, underscoring the clinical viability of the method by avoiding excessive false alarms under more challenging conditions.

Overall, although absolute numbers vary across datasets due to differences in experimental protocols and dataset characteristics, the results demonstrate that the proposed framework can be effectively re-optimized to achieve competitive performance in multiple independent datasets. This adaptability is crucial for future clinical deployment, as it suggests that the same methodological backbone can be tailored to new datasets without redesign, providing a reproducible and reliable reference point for further FoG prediction research.

## V. Discussion

The present results indicate that reliable advance warning of freezing of gait can be sustained within a clinically meaningful time window. Under a horizon-conditioned evaluation, performance on our clinical cohort remains stable up to roughly 2 s before onset and then declines as the evaluation horizon increases. We interpret this curve as a quantitative budget that, for any operating point, the profile specifies how much lead time is available at a given reliability after accounting for sensing, inference, communication, and user response. Framed this way, the analysis separates near-onset detection from genuine forecasting and offers a common foundation for comparing design choices such as windowing, encoder, and post-processing without conflating onset-adjacent evidence with earlier cues.

Crucially, this capacity for advance warning appears robust to disease progression. Our subject-level analysis revealed that prediction performance was not statistically correlated with higher FOGQ scores, age, or disease duration. This dissociation implies that the derived horizon profiles represent a generalized capability applicable even to patients with severe motor impairment, rather than being an artifact of testing on milder cases.

Beyond this framing, the study strengthens evidence by grounding the analysis in a larger clinical cohort of 55 patients and by validating, with harmonized preprocessing, on two public benchmarks (Daphnet [22] and BXHC [23]). Taken together, the combination of horizon-aware reporting, a larger cohort, and cross-dataset validation provides a reproducible basis for quantifying usable advance time rather than only reporting pooled window scores.

Several factors bound the interpretation and generality of the findings. First, recordings were obtained in structured, supervised tasks rather than in free-living conditions. Daily life introduces distractions, dual-task demands, irregular terrains, and variable walking speeds. These context changes can alter pre-onset signatures and shift the operating point along the horizon curve. Establishing external validity will require prolonged ambulatory monitoring with reliable event annotation.

Second, the intrinsic data regime imposes constraints that no evaluation protocol can fully eliminate. Pre-FoG segments are short and sparse compared with normal gait and FoG, which amplifies class imbalance and concentrates errors in the pre-FoG class that matters for prevention. As the evaluation horizon increases and onset-adjacent windows are withheld, the number of positive samples decreases, and the uncertainty of long-horizon estimates grows. This effect is visible even when overall metrics are high and should be taken into account when interpreting early-warning capability.

Third, labeling necessarily abstracts a gradual transition. A fixed pre-FoG region, selected through model-selection procedures as Bayesian Optimization, provides a practical default for training. Nevertheless, the true span of pre-onset degradation likely varies across patients and even across episodes for the same patient. This variability can blur decision boundaries and partially explain gaps between strong pooled performance and weaker precision at earlier horizons. Exploring patient- or episode-adaptive labeling schemes, including soft labels that encode uncertainty near class boundaries, may improve alignment between supervision and physiology.

Fourth, although preprocessing was harmonized within our experiments, absolute performance values remain dataset-specific because training and testing were conducted separately on each dataset to verify the validity of the proposed evaluation protocol. This design isolates dataset effects and ensures that observed horizon–performance trends are not confounded by distributional differences. However, future studies should explore joint or multi-dataset training strategies that integrate complementary information across cohorts. Such approaches could enhance statistical power, improve model robustness, and provide a more comprehensive view of the temporal dynamics that precede freezing events.

Finally, the present analysis focuses on inertial sensing. Additional modalities such as surface EMG or contextual metadata might disambiguate subtle pre-onset patterns and reduce false alarms at longer horizons, yet they introduce tradeoffs in wearability, annotation effort, and power consumption. The extent to which such information improves early-horizon reliability without compromising practicality remains an open empirical question.

Building on the limitations outlined above, several methodological directions follow naturally. First, one possible direction is to encourage wider adoption of horizon-aware reporting, so that results can be interpreted in terms of usable lead time rather than a single pooled number. Such practice would help distinguish improvements that rely on onset-adjacent evidence from those that remain stable earlier in time. Second, learning process should be made more resilient to scarcity and ambiguity in pre-FoG data. Training schedules that place greater emphasis on earlier horizons and loss functions that counter class imbalance can stabilize precision where data are scarcest, while patient- or episode-adaptive labeling potentially with soft targets near the transition may better reflect the gradual transition into freezing.

A complementary pathway is to broaden the empirical basis of evaluation. Although we trained and tested separately on each cohort to isolate dataset effects and verify the protocol, integrating cohorts through joint or multi-dataset training could increase statistical power, improve robustness, and reveal common temporal structure preceding freezing. In parallel, extending validation to longer free-living recordings and pairing window-level horizon profiles with episode-level summaries would align evaluation more closely with operational goals, providing a clearer view of when predictions translate into actionable early warnings.

## VI. Conclusion

This study presented a Transformer-based framework for freezing of gait prediction that couples architectural design and data preprocessing with a progressive-learning strategy, jointly tuned using Bayesian optimization. Evaluations on a 55-patient clinical dataset and two public datasets demonstrate that, under Freezing Prediction Horizon evaluation, performance remains stable up to approximately 2 s before FoG onset on our dataset and then gradually declines. Crucially, subject-level analysis indicates that this predictive capability is robust to disease severity, suggesting that the established horizon profile is applicable across varying degrees of motor impairment. The resulting horizon–performance profile complements conventional patient-dependent metrics by quantifying achievable advance time at specified reliability scores. From a prevention perspective, this evaluation offers a reproducible and interpretable summary of early-warning capacity at specified accuracy levels and may guide design targets when combined with system latency characterization. Overall, horizon-conditioned evaluation reframes FoG prediction around clinically actionable lead time and represents a step toward translating algorithmic reliability into real-world, prevention-oriented applications. Future work will extend validation to free-living settings, examine robustness across datasets and protocols, and explore adaptive, multimodal approaches for more dependable early prediction in diverse patient populations.

## Data Availability

The clinical dataset analyzed in this study contains human participant data and is not publicly available due to privacy and ethical restrictions. De-identified data may be made available from the corresponding authors upon reasonable request and subject to institutional approval and a data use agreement. The public datasets used in this study are available from their respective sources.

## Appendix I

Bayesian optimization Analysis

### A. Search Space and Optimal Configuration

Bayesian optimization (BO) was employed to jointly explore data parameters (pre-FoG region, window size) and model hyperparameters in order to avoid arbitrary design choices. Configurations were evaluated, balancing model depth, attention mechanisms, feed-forward dimensionality, and training parameters. The complete search space and the best-performing configuration are summarised in Table IV.

The optimization process revealed that performance is jointly shaped by data representation and model capacity. In particular, longer pre-FoG regions consistently allowed higher potential scores across folds, while smaller regions restricted the model to less informative temporal context.

Similarly, intermediate network depths achieved more stable convergence compared to either very shallow or excessively deep architectures, reflecting the trade-off between expressive power and overfitting risk on relatively small patient datasets. The final selected configuration consists of 4 encoder blocks, 2 attention heads, a model dimension of 256, a feed-forward expansion factor of 7, dropout of 0.25, and a learning rate of 9.5 × 10^*−*4^. This setup, combined with a 5 s pre-FoG region and 1 s window size, was adopted for all subsequent experiments. Beyond its competitive accuracy, this configuration was chosen because it balances predictive power with computational efficiency, ensuring feasibility for downstream applications such as wearable device deployment.

### B. Effect of pre-FoG Region Length

Figure 8 (top row) compares four candidate pre-FoG region lengths (2–5 s). While median Macro F1 values remain comparable across settings, the distribution of outcomes indicates that longer regions, particularly 5 s, enable substantially higher maximum scores. This behaviour reflects two key mechanisms: (i) additional temporal context improves the model’s ability to capture gradual degradation of gait preceding a freeze; and (ii) longer pre-FoG regions increase the number of positive pre-FoG samples, mitigating the class imbalance that otherwise hampers training.

**Fig. 8:**
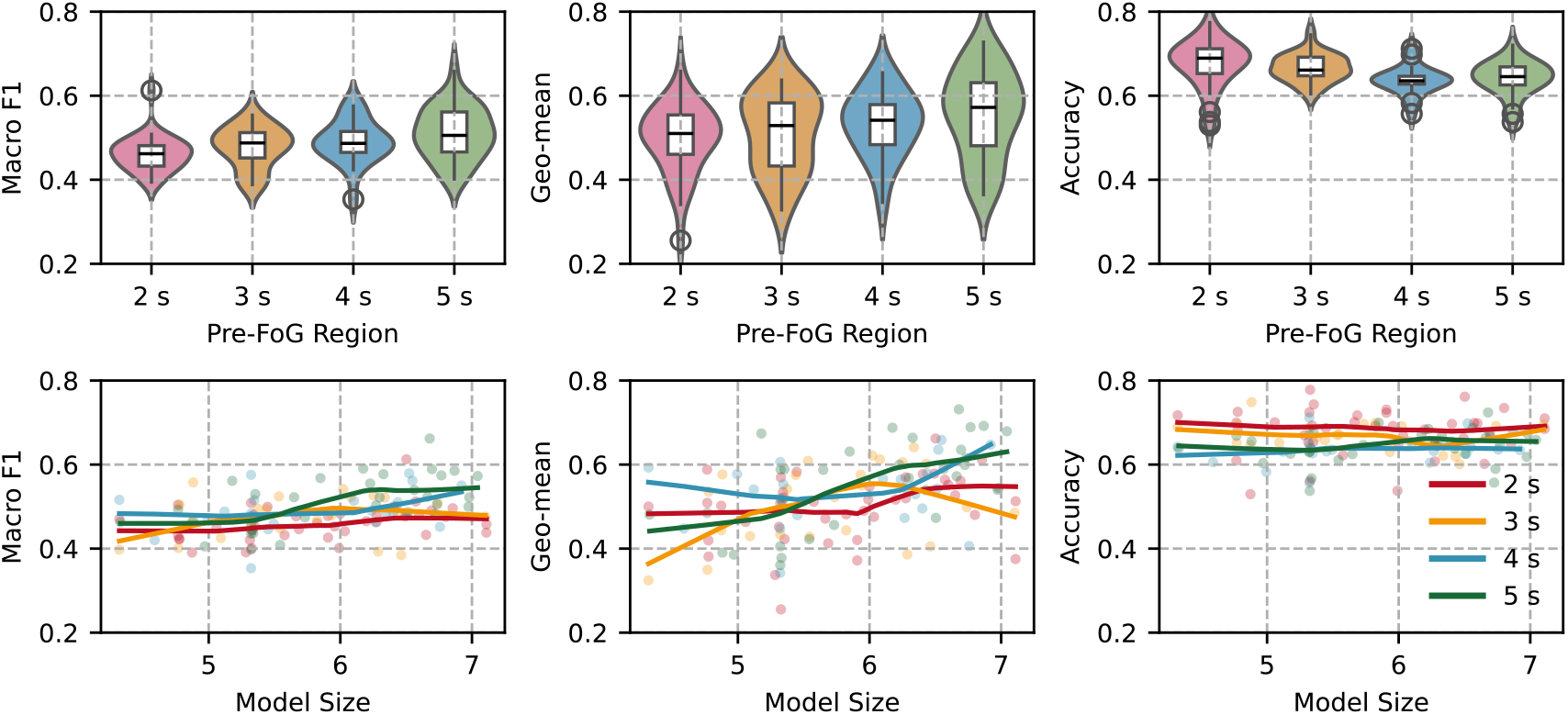
Influence of pre-FoG region length on three key metrics. The top row shows violin plots for Macro F1, Geo-mean and Accuracy obtained from Bayesian-optimised models, grouped by pre-FoG region length (2–5 s). The bottom row depicts LOWESS fits (solid lines) of each metric against model size (MB, log scale on the *x*-axis).

These results support the decision to adopt 5 s as the default pre-FoG region in subsequent experiments. Importantly, the analysis also confirms that using excessively short regions (e.g., 2 s) risks discarding informative gait dynamics and constraining model generalizability. By contrast, 5 s strikes a balance between providing enough context and maintaining realistic clinical lead times for intervention.

### C. Model Complexity and Predictive Performance

Figure 8 (bottom row) and Figure 9 examine the link between model size and predictive performance. At short pre-FoG durations (2–4 s), Spearman correlations between model size and Macro F1 were weak to moderate (| *ρ*| ≤ 0.37), suggesting that limited context constrains the benefit of larger architectures. However, at 5 s, correlation strength increased markedly (|*ρ*| = 0.57 for both Macro F1 and Geo-mean), indicating that when sufficient temporal information is available, larger models can more effectively exploit their capacity to detect subtle pre-FoG patterns. This trend highlights an important design implication: model architecture and data representation must be optimized jointly. Increasing model size alone does not guarantee performance gains unless adequate temporal context is provided. Conversely, longer pre-FoG windows without sufficient model complexity may leave predictive patterns underutilised. The consistently weak correlation with Accuracy (|*ρ*| ≤ 0.23) further underscores the inadequacy of Accuracy in imbalanced FoG prediction, reinforcing the need for Macro F1 as the primary evaluation metric.

**Fig. 9:**
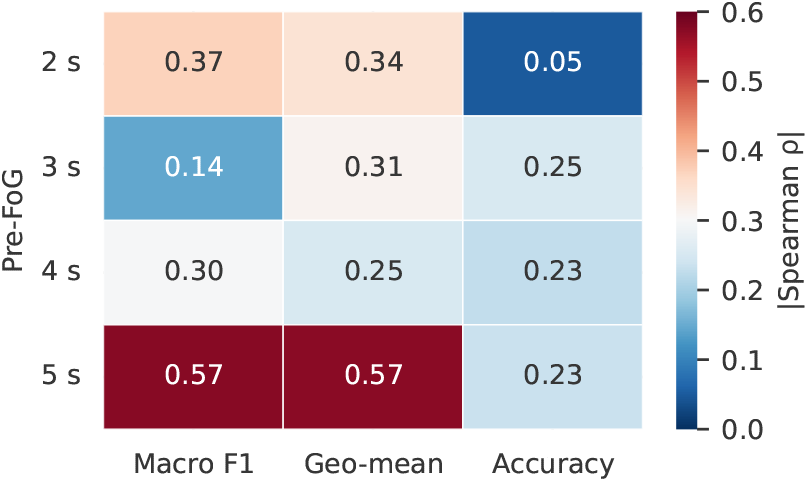
Absolute Spearman correlation (|*ρ*|) between model size (base–10 logarithm of trainable parameters) and three evaluation metrics for each pre-FoG region. Darker shades indicate stronger correlations. A 5s pre-FoG region exhibits the highest association between model capacity and both Macro-F1 and Geo-mean, whereas Accuracy is largely insensitive to model size.

Together, these findings provide a principled justification for our final configuration, which leverages both a sufficiently long pre-FoG region and a moderately deep Transformer architecture.

## Appendix II

Additional Quantitative Results

Detailed numerical values corresponding to Figures 3 and 7 are provided in Tables V and VI, respectively.

Table V reports the class-wise and overall results of the patient-dependent (PD) evaluation on our clinical dataset, showing mean ± standard error (SE) over 55 subjects. Table VI presents cross-dataset results under the Stratified K-Fold evaluation for both the Daphnet and BXHC datasets, including all metrics used in prior studies to ensure consistent benchmarking.

**TABLE V:** Detailed patient dependent results on our clinical dataset. Values represent the mean ± standard error across 55 subjects, reported for each gait class as well as the overall scores. These statistics provide a numerical reference for Figure 3, showing inter-class variability and the relative contribution of Pre-FoG detection to overall model performance.

| Class | Accuracy | Precision | Sensitivity | Specificity | Geo-mean | Macro F1 |
| --- | --- | --- | --- | --- | --- | --- |
| Normal gait | 92.12 ± 0.69 | 90.77 ± 0.95 | 90.20 ± 1.08 | 86.90 ± 2.10 | 87.16 ± 1.81 | 90.21 ± 1.00 |
| Pre-FoG | 92.28 ± 0.63 | 79.58 ± 1.44 | 73.58 ± 1.90 | 95.42 ± 0.51 | 82.86 ± 1.20 | 75.07 ± 1.63 |
| FoG | 94.83 ± 0.46 | 86.43 ± 1.55 | 86.24 ± 1.97 | 95.08 ± 0.64 | 89.15 ± 1.41 | 85.35 ± 1.84 |
| Macro / Overall | 89.61 ± 0.83 | 85.53 ± 0.85 | 83.34 ± 1.04 | 91.25 ± 1.76 | 85.17 ± 1.82 | 83.49 ± 0.96 |

**TABLE VI:**
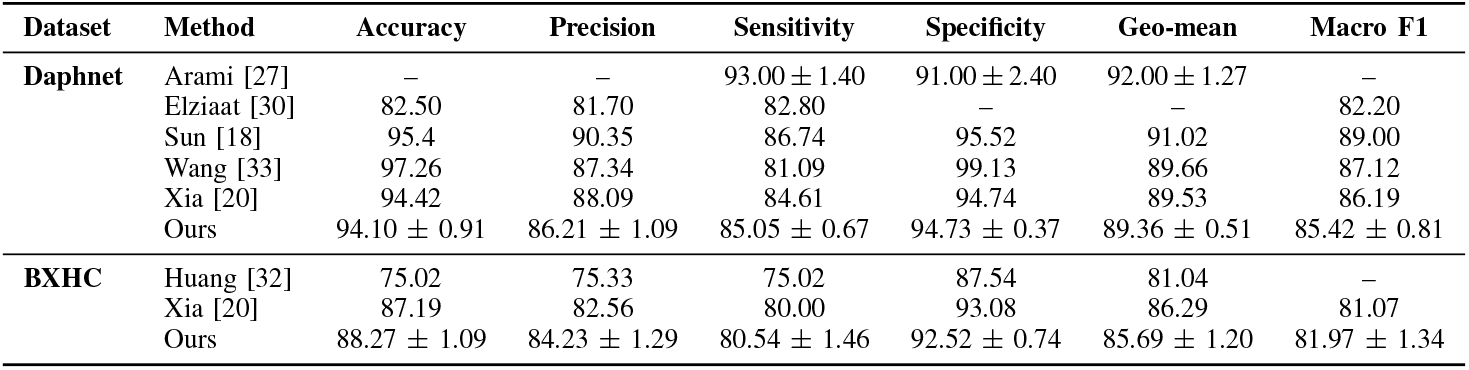
Numerical comparison of patient dependent performance under the Stratified K-Fold evaluation across datasets. The table lists mean ± standard error values for all available metrics, enabling direct benchmarking between prior works and the proposed model on the Daphnet and BXHC datasets. For Arami [27], the SE values were estimated from the reported standard deviation to maintain consistency of notation. This table complements Figure 7 by providing exact quantitative results for reproducibility and cross-study comparison.

| Dataset | Method | Accuracy | Precision | Sensitivity | Specificity | Geo-mean | Macro F1 |
| --- | --- | --- | --- | --- | --- | --- | --- |
| **Daphnet** | Arami [27] | – | – | 93.00 ± 1.40 | 91.00 ± 2.40 | 92.00 ± 1.27 | – |
|  | Elziaat [30] | 82.50 | 81.70 | 82.80 | – | – | 82.20 |
|  | Sun [18] | 95.4 | 90.35 | 86.74 | 95.52 | 91.02 | 89.00 |
|  | Wang [33] | 97.26 | 87.34 | 81.09 | 99.13 | 89.66 | 87.12 |
|  | Xia [20] | 94.42 | 88.09 | 84.61 | 94.74 | 89.53 | 86.19 |
| | Ours | 94.10 ± 0.91 | 86.21 ± 1.09 | 85.05 ± 0.67 | 94.73 ± 0.37 | 89.36 ± 0.51 | 85.42 ± 0.81 |
| **BXHC** | Huang [32] | 75.02 | 75.33 | 75.02 | 87.54 | 81.04 | – |
|  | Xia [20] | 87.19 | 82.56 | 80.00 | 93.08 | 86.29 | 81.07 |
| | Ours | 88.27 ± 1.09 | 84.23 ± 1.29 | 80.54 ± 1.46 | 92.52 ± 0.74 | 85.69 ± 1.20 | 81.97 ± 1.34 |

Together, these tables provide a comprehensive reference for reproducibility and numerical verification of the reported results, highlighting that our proposed model maintains balanced performance across datasets and evaluation protocols.

## Notes

* This work was supported in part by NUS-NNI 2016 Grant R263000C36133 and in part by NMRC/CISSP/2014/2015.

### Competing Interest Statement

The authors have declared no competing interest.

### Funding Statement

This work was supported in part by the NUS-NNI 2016 Grant R263000C36133 and in part by the National Medical Research Council, Singapore (NMRC/CISSP/2014/2015).

### Author Declarations

Ethical approval was obtained from the SingHealth Centralised Institutional Review Board (CIRB Ref: 2016/2743; approved on 28 September 2016). Data collection was conducted at two clinics of the National Neuroscience Institute, Singapore, from September 2016 to August 2017.

